# Risk factors for developing symptomatic COVID-19 in older residents of nursing homes: A hypothesis-generating observational study

**DOI:** 10.1101/2022.01.18.22269433

**Authors:** Anna Escribà-Salvans, Sandra Rierola-Fochs, Pau Farrés-Godayol, Miriam Molas-Tuneu, Dyego Leandro Bezerra de Souza, Dawn A Skelton, Ester Goutan-Roura, Daniel Alonso Masmitjà, Eduard Minobes-Molina, Javier Jerez-Roig

## Abstract

**Background:** COVID-19 pandemic has had a major impact on society, including on residents of nursing homes (NH), who have a higher risk of complications and mortality due their physical and intellectual disabilities.

**Aim:** To identify which risk factors associated with developing COVID-19 infection with symptoms in institutionalized older people.

**Methods:** A 1-year longitudinal multicenter study was conducted in 5 NH during the period December 2019 to March 2021. The inclusion criteria used were residents aged 65 years or over, living in the NH permanently, with a diagnostic test for COVID-19 confirmed by reverse transcription polymerase chain reaction and/or serological test. The main variable was symptomatic COVID-19, with at least one of the following symptoms (fever, respiratory difficulties, cough, diarrhea, sudden urinary incontinence and disorientation or delirium). Three assessments were performed: baseline, six and twelve months follow-up. Descriptive and bivariate analysis (calculating relative risk-RR) were performed, considering a 95% confidence level and a statistically significant *p* <0.05.

**Results:** Of the total sample of 78 individuals who tested positive for COVID-19, mean age 84.6 years (SD=±7.8), 62 (79.5%) were female; 40 (51.3%) participants presented with COVID-19 symptoms. Living in a private NH (RR=3.6, 95% CI [1.2–11.0], *p*=0.023) and having suffered a stroke (RR=4.1, 95% CI [1.1–14.7], *p*=0.033) were positively associated with developing COVID-19 infection with symptoms.

**Conclusions:** Having suffered a stroke and living permanently in a private health care facility were positively associated with symptomatic COVID-19 in this sample of institutionalized older people.

## INTRODUCTION

In December 2019, the COVID-19 pandemic, started to have a major impact on society^1^. COVID-19 or severe acute respiratory syndrome coronavirus 2 (SARS-COV-2) has had a continuing unprecedented effect on worldwide health and economic systems^2^. It is estimated that 69% of deaths due to COVID-19 in Spain between April and June 2020 were in older people living in nursing homes (NHs)^3^.

The main symptoms of COVID-19 are fever, respiratory difficulties, cough, diarrhea, sudden urinary incontinence (UI) and disorientation or delirium^4^. Fever, defined as a temperature above 37.5°, is one of the most common symptoms in COVID-19 viral infection^5^. Respiratory distress and cough, defined as increased respiratory rate and the need to keep the airway clear due to difficulty^5^. Gastrointestinal symptoms, such as diarrhea along with other gastrointestinal symptoms, were defined in the context of COVID-19 infection as intestinal distress: the main symptom was diarrhea followed by nausea, vomiting and anorexia^5^. Sudden UI is a less frequent symptom in COVID-19 positive patients and difficult to make a differential diagnosis in institutionalized older people^6^. However, studies present clinical suspicion that COVID-19 infection leads to viral cystitis causing an increase in urinary frequency with the onset of sudden UI^6^. Finally, delirium and disorientation due to COVID-19 are understood as fluctuating alteration of consciousness, inattention and reduced awareness. Some patients may show hallucinations^7^.

One of the defining characteristics of COVID-19 was its severity within older adults and frailer multimorbid younger adults. They were much more prone to become critically ill with disorders of atypical symptoms and multiple organ abnormalities. They were also much more likely to die of their infection^7^. There are multiple risk factors associated with higher risk of mortality: aging, male sex, hospital admission required for COVID-19, having been hospitalized in the month before the admission for COVID-19, underlying diseases like hypertension, cardiovascular disease, dementia, respiratory disease, liver disease, diabetes with organ damage, cancer, chronic obstructive pulmonary disease, chronic renal disease, malnutrition, immunodeficiency, specific genotypes of interleukins and interferons, asthma, autoimmune diseases such as multiple sclerosis, rheumatoid arthritis and systemic lupus erythematous, cerebrovascular disease and chronic liver disease^8^.

Older people who reside in NHs, therefore, had a higher risk of complications and mortality from COVID-19 infection due to their more advanced age, greater number of underlying chronic conditions, cognitive and physical impairments and frailty than the rest of the older population^4,8^. Environmental factors must also be taken into account: space, inadequate infrastructure and difficulty in maintaining social distance between residents and NH staff all increased the risk of infection. Nursing homes are particularly fragile institutions when dealing with an infectious disease outbreak, such as influenza virus, norovirus or the current COVID-19^9^. In Catalonia, from the beginning of the pandemic until June 2020, 3965 people died from various causes in NHs. Of these, 1133 institutionalized older persons died from COVID-19 in private NHs and 222 in public NHs^3^.

Even before the COVID-19 pandemic, Fité et al., (2019)^10^, reported that the staffing level and health of Catalan NHs personnel was precarious: low salaries, lack of trained personnel and at the physical and psychological limit due to working conditions^10^. With the COVID-19 outbreak, the staffing situation worsened further, being exposed to high levels of workload and social pressure^11^. Some studies^9,11^ indicates that factors such as lack of staff due to COVID-19 infection, their scarce training in the management of infectious diseases or in the correct use of personal protective equipment (PPE), insufficient foresight and contingency planning due to lack of coordination of health and social services and NHs, provided the perfect environment for COVID-19 to spread rapidly among residents and staff of NHs.

To our knowledge, this is the first study to collect sociodemographic and health-related data just prior to the onset of the COVID-19 pandemic, to identify which are the risk factors of developing COVID-19 infection with symptoms in institutionalized older people. This information may be useful to better understand the causality of COVID-19 occurrences and thus be able to design strategies aimed at its prevention in the future.

## METHODOLOGY

### Study design

An observational hypothesis-generating design was conducted in an one year cohort longitudinal multicentre study (Clinical Trials ID: NCT04297904) that follows the standards of STROBE (Strengthening the Reporting of Observational studies in Epidemiology) for cohort studies^12^. It is a sub-study from the OsoNaH project^13^ and baseline data collection was conducted between December 2019 and March 2020 (before the state of alarm in Spain due the COVID-19 pandemic). The study was carried out in five NHs of Osona (a region of Central Catalonia, Spain).

### Participants

All geriatric residents aged 65 years or over living in the NH permanently were included. Residents in a coma or in palliative care (short-term prognosis) and those who refused (or their legal guardian) to participate in the study, were excluded. In addition, those who did not have positive COVID-19 diagnosis and those who did not perform any COVID-19 diagnostic test were also excluded in this sub-analysis.

### Consent and ethical approval

Ethical permission was obtained by the Ethics and Research Committee of the University of Vic - Central University of Catalonia (registration number 92/2019 and 109/2020). Signed informed consent was gained from the resident or his/her legal guardian. The project meets the criteria required in the Helsinki Declaration, as well as the Organic Law 3/2018 (December 5) on the Protection of Personal Data and Guarantee of Digital Rights.

### Study procedures

Before starting data collection, every NH director accepted the participation in the project with a formal consent. After that, the list of residents was obtained, and the individuals were selected according to inclusion/exclusion criteria. Then, the residents or their legal guardians were informed about the project and those who accepted to participate signed the informed consent. At the beginning of the project, the research team collecting the data was trained in the use of all tools and tests. The reliability of the data collected through the health and sociodemographic questionnaires was evaluated by calculating the Kappa index and the interclass correlation coefficient (ICC) for the data of 20 residents. The CCI scores were greater than 0.75 on all physical tests. The results of these 20 residents were included in the final total study sample.

### Outcome measurements

The main outcome, symptomatic COVID-19 infection, was considered if the resident tested positive for reverse transcription polymerase chain reaction (RT-qPCR) and/or serologic testing during the 1-year follow-up period and experienced the presence of at least one COVID-19-related symptom (fever, respiratory distress, cough, diarrhea, sudden UI, and disorientation or delirium)^4^. Evidence showed that more than 50% of COVID-19 positive residents could be asymptomatic. Therefore, we performed a RT-qPCR testing on all residents, whether or not they were symptomatic, following the same strategies as Lopez de la Iglesia^14^.

The presence of at least one COVID-19-related symptom with a positive COVID-19 test, the primary variable, was collected in two waves: one six months after the baseline and the other one year after the baseline.

The rest of sociodemographic and clinical outcomes were collected at baseline, at the start of the study in December 2019 just prior to the onset of the COVID-19 pandemic. Sociodemographic information and medical history, such as age, sex, education level, marital status, children, chronic diseases, smoking and drinking habits, type of NH and hospitalizations, was obtained from NH records and checked with NH professionals. Anthropometric variables such as body mass index (BMI), weight and height were measured with a Seca 213 measuring device, the Tanita TBF-300. Nutritional status was assessed using the Mini Nutritional Assessment (MNA)^15^. Continence status was reported using section H of the Minimum Data Set (MDS) version 3.024^16^. Functional capacity was measured using the modified Barthel Index, excluding continence items^17^. Cognitive status was assessed using the Pfeiffer Scale^18^. The SARC-F questionnaire was used to determine the risk of developing sarcopenia^19^. Sedentary behaviour (SB) was assessed by the activPAL3TM activity monitor (PAL Technologies Ltd., Glasgow, UK), considered the gold standard on assessing SB and postural changes in older adults^20^. The device was placed in the middle of the right thigh or in the non-plegic thigh in subjects with stroke and worn for 7 consecutive days^21^. The following variables were extracted from the device: waking hours mean, steps per day mean, average time in hours of upright position, transitions of sit to stand per day mean, average time in hours of sitting, percentage of time sitting in waking hours and average time of SB bouts in minutes. Frailty was assessed using the Clinical Frailty Scale (CFS)^22^. Physical capacity was examined using the Short Physical Performance Battery (SPPB)^23^. The authors received permission to use all of these instruments from the copyright holders. The approximate time to complete the physical tests and questionnaires with each resident was 30-45 minutes. Further details, please refer to the Clinical Trials protocol (Clinical Trails ID: NCT04297904) and Osonah Project^13^.

### Statistical analysis

A descriptive analysis was performed indicating absolute and relative frequencies for categorical variables. Bivariate analysis was applied using the Chi-square test (or Fisher’s test, when necessary) and the linear Chi-square test for dichotomous and ordinal variables, respectively. As a measure of association, Relative Risk (RR) was calculated, with a confidence level of 95%. Multivariate analysis was performed by linear logistic regression with robust variance. A level of *p* < 0.05 was statistically significant. Data were analyzed with SPSS version 27 (SPSS Inc., Chicago IL).

## RESULTS

The initial sample of the study was 185 participants, 53 (28.6%) were excluded as they did not meet the inclusion criteria at baseline. After 12 months follow up, 54 (29.2%) had not been tested as COVID-19 positive or did not perform any diagnostic test so were excluded. A total of 78 residents, 62 (79.5%) of whom were women, representing 41.2% of the total number of initially recruited residents in the five NHs were included (Fig 1).

**Fig 1.**
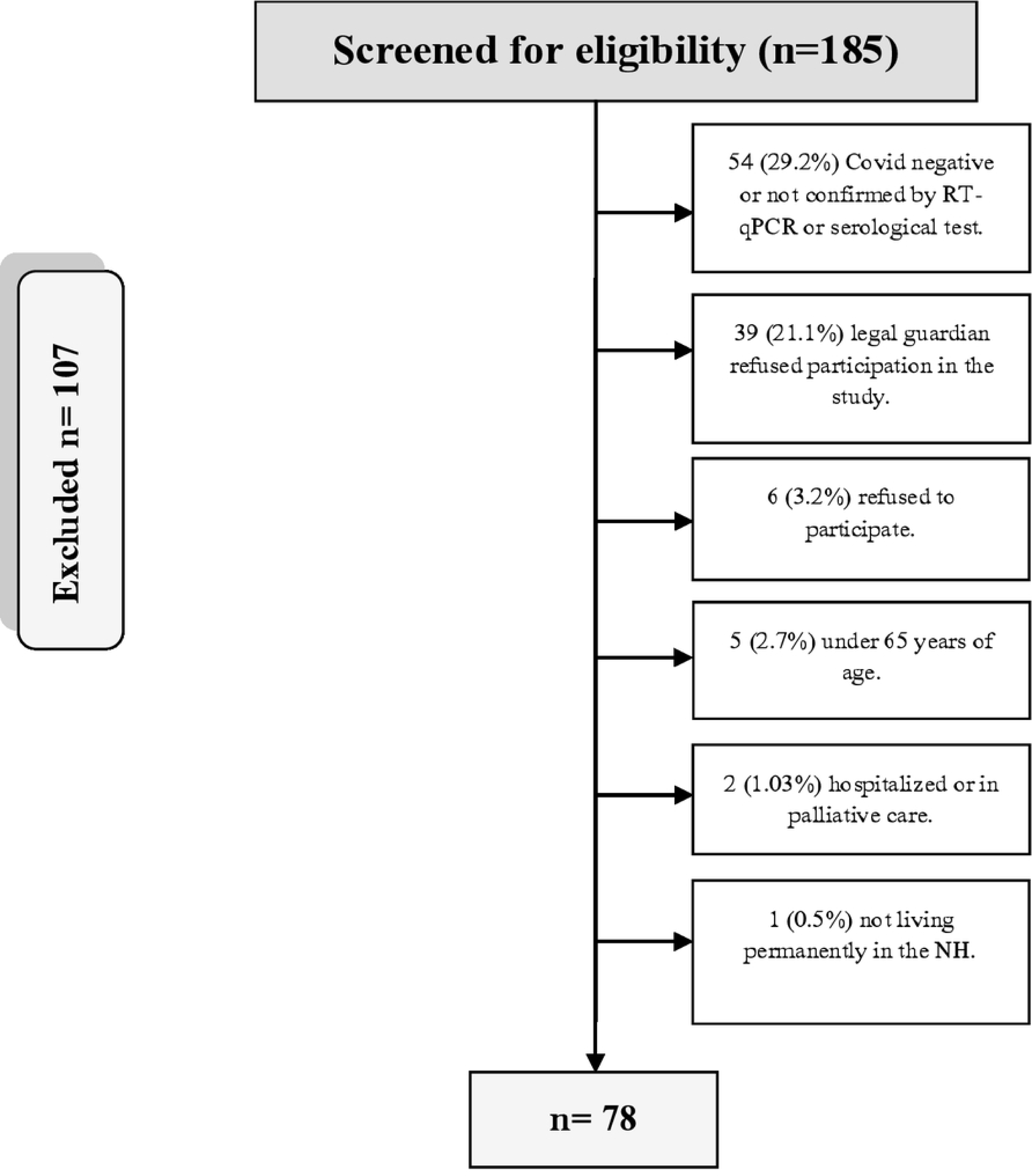
Flow chart of the sampling process

The mean age of the participants was 84.6 years (SD=7.8) of which 46 (59%) were over 86 years old. Fifty-nine (75.7%) were single or widowed. In terms of offspring, 61 (78.2%) had one or more children. Four (5.1%) were smokers and 9 (11.6%) were alcohol consumers. Thirty-eight (48.7%) had been living permanently in the NH for more than two years. Fifty-one (65.4%) lived in a subsidized NH and 27 (34.6%) in a private NH.

### Chronic Pathologies

Forty-three (55.1%) participants had pluripathology (more than 5 diagnosed chronic pathologies). The most relevant chronic conditions were: hypertension (64.1%), dementia (52,6%), cardiac pathology (43.6%) and previous stroke (19.2%).

### Intellectual, Continence, Nutritional and Physical Conditions

Cognitively, 55 (70.5%) geriatric residents had moderate or severe cognitive impairment. Twenty-eight (35.9%) participants had urinary incontinence and 14 (17.9%) faecal incontinence. At nutritional level, 11 (14.1%) geriatric residents lost weight in the last year, 25 (32.1%) were malnourished or at risk of malnutrition and 46 (59.0%) were obese. Regarding functional capacity, 62 (79.5%) had moderate or severe dependence. Thirty-five (44.9%) had risk of sarcopenia. Forty (51.3%) had moderate, severe or extreme frailty. At physical performance level, 40 (51.3%) were disabled.

### Sedentary Behaviour Pattern

They performed an average of 11.1 hours (SD: 1.1) waking hours; they walked an average of 1599.0 (SD: 2551.0) steps per day, had an average of 1.9 (SD: 1.9) hours in an upright position and 19.0 (SD: 14.0) transitions of sit to stand per day. On the other hand, the residents spent a mean of 9.1 (SD: 1.6) hours in SB, which means an average of 82.6% (SD: 16.4) of their waking hours spent in SB. The average SB bout was 62.3 (SD: 64.5) minutes.

### Follow-up

At twelve months, 40 (21.6%) deaths were registered, of which 19 (10.3%) were COVID-19 deaths. One (0.5%) person in the study was not followed up as she moved out of the NH. Forty individuals (51.3%) of the total sample of residents who tested positive had COVID-19 symptoms.

The rest of the sociodemographic and health conditions of institutionalized older adults with a confirmed diagnosis of COVID-19 are shown in Table 1.

**Table 1.** Descriptive analysis of the sample of institutionalized older adults with confirmed diagnosis of COVID-19 (n=78)

| Variables | Frequency (%) / Mean (SD) |
| --- | --- |
| **Education level** |  |
| Illiterate | 19 (24.4) |
| Primary education | 34 (43.6) |
| Secondary education/ University studies | 25 (32.0) |
| **Marital Status** |  |
| Married/ Partner | 12 (15.4) |
| Single | 12 (15.4) |
| Widowed | 47 (60.2) |
| NR | 7 (9.0) |
| **Children** |  |
| 0 | 17 (21.8) |
| 1 | 20 (25.6) |
| +2 | 41 (52.6) |
| **Chronic diseases:** |  |
| Hypertension | 50 (64.1) |
| Dementia | 41 (52.6) |
| Cardiac pathology | 34 (43.6) |
| Dyslipidemia | 22 (28.2) |
| Kidney failure | 21 (26.9) |
| Diabetes mellitus | 19 (24.4) |
| Depression | 18 (23.1) |
| Stroke | 17 (21.8) |
| Digestive pathology | 16 (20.5) |
| Cancer | 15 (19.2) |
| Mental pathology | 15 (19.2) |
| Osteoporosis | 15 (19.2) |
| Pulmonary pathology | 14 (17.9) |
| Hypothyroidism | 12 (15.4) |
| Circulatory pathology | 12 (15.4) |
| Parkinson | 10 (12.8) |
| Anaemia | 9 (11.5) |
| **Hospitalizations** | 16 (20.5) |
| **Functional capacity (Barthel Index)** |  |
| Independence | 4 (5.1) |
| Slight dependence | 12 (15.4) |
| Moderate dependence | 29 (37.2) |
| Severe dependence | 33 (42.3) |
| **Cognitive status (Pfeiffer Scale)** |  |
| No cognitive impairment | 8 (10.3) |
| Mild cognitive impairment | 13 (16.7) |
| Moderate cognitive impairment | 20 (25.6) |
| Sever cognitive impairment | 35 (44.9) |
| NR | 2 (2.5) |
| **Physical capacity (SPPB)** |  |
| Robust person | 5 (6.4) |
| Pre frail person | 10 (12.8) |
| Frail person | 19 (24.4) |
| Disabled person | 40 (51.3) |
| NR | 4 (5.1) |
| **Frailty (CFS)** |  |
| Excellent/ Acceptable/ Good | 17 (21.8) |
| Vulnerable | 6 (7.7) |
| Minimal frailty | 13 (16.7) |
| Moderate frailty | 17 (21.8) |
| Severe/ Extreme frailty | 23 (29.5) |
| Terminal patient | - |
| NR | 2 (2.5) |

SD: Standard Desviation; SPPB: Short Physical Performance Battery; CFS: Clinical Frailty Scale; NR: Not reported due to COVID19 pandemic outbreak

### Associations

In the bivariate analysis, symptomatic cases of COVID-19 were significantly associated with living in a private residence (*p*=0.008) and having had a stroke (*p*=0.019). The rest of the sociodemographic and health variables showed no significance (Table 2). In the multivariate analysis, living in a private residence and having had a stroke were positively associated with having symptomatic COVID-19, independently of age and sex. The final model was adjusted according to the Hosmer Lemeshow test (*p*=0.933).

**Table 2.**
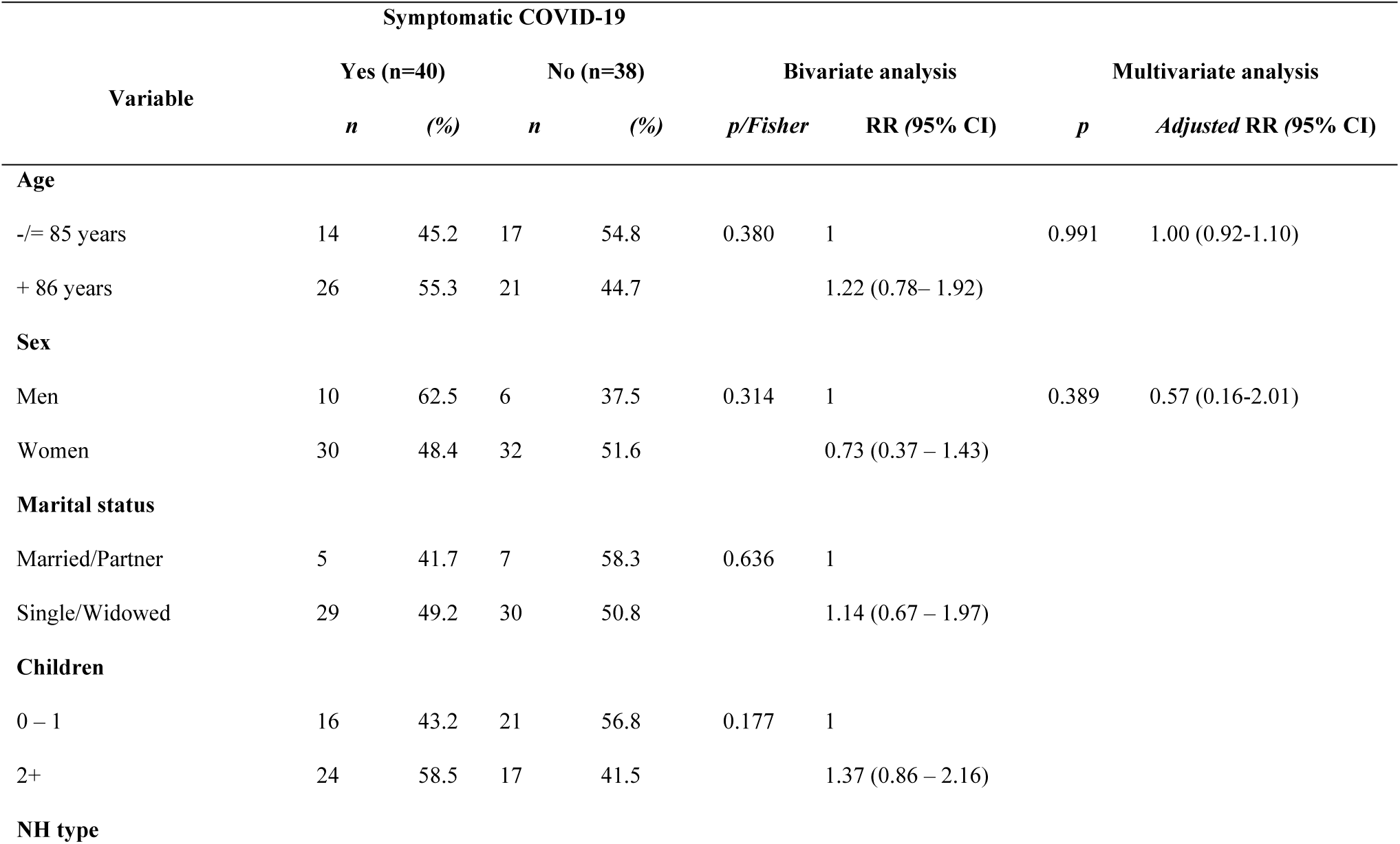
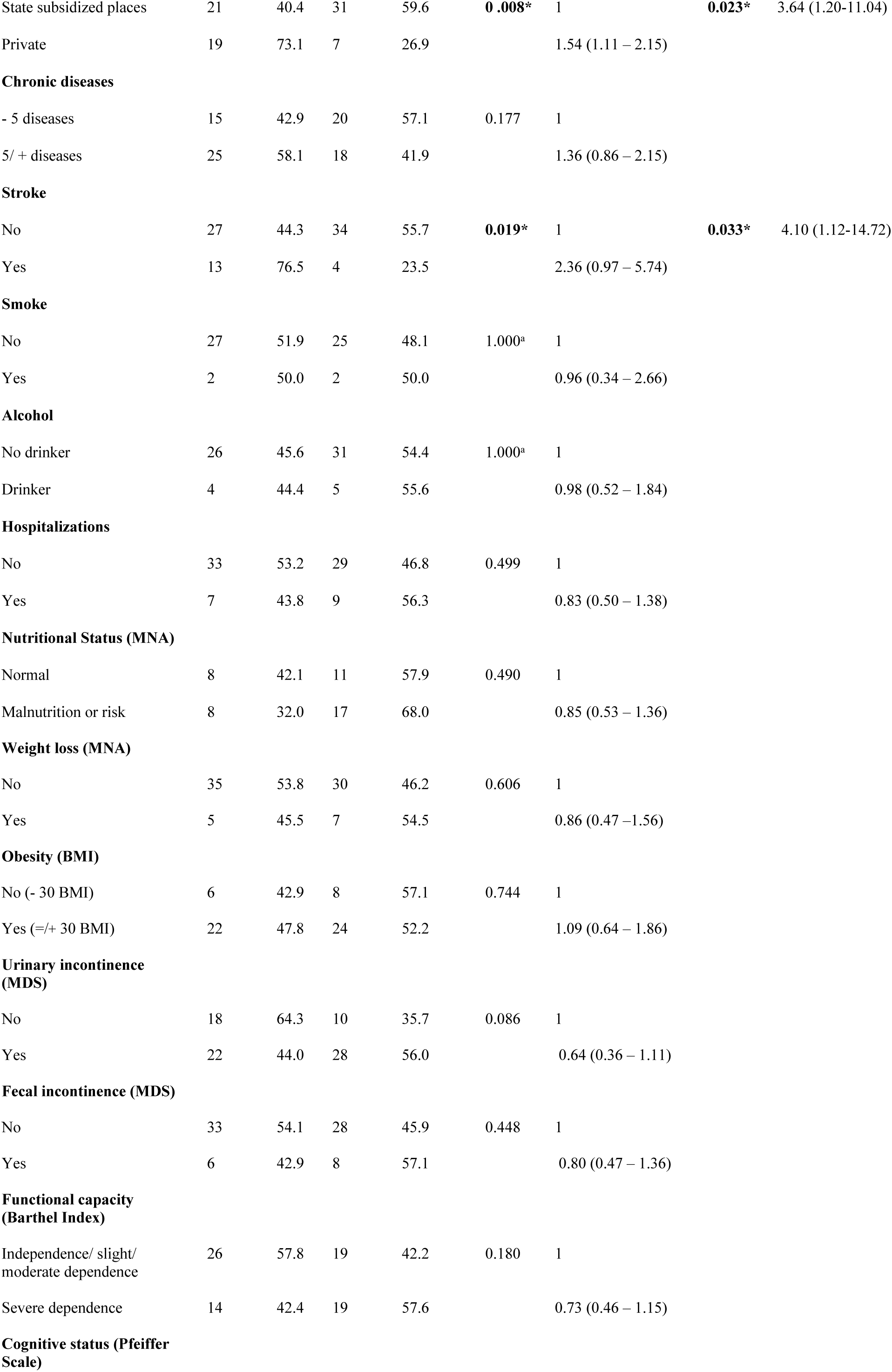
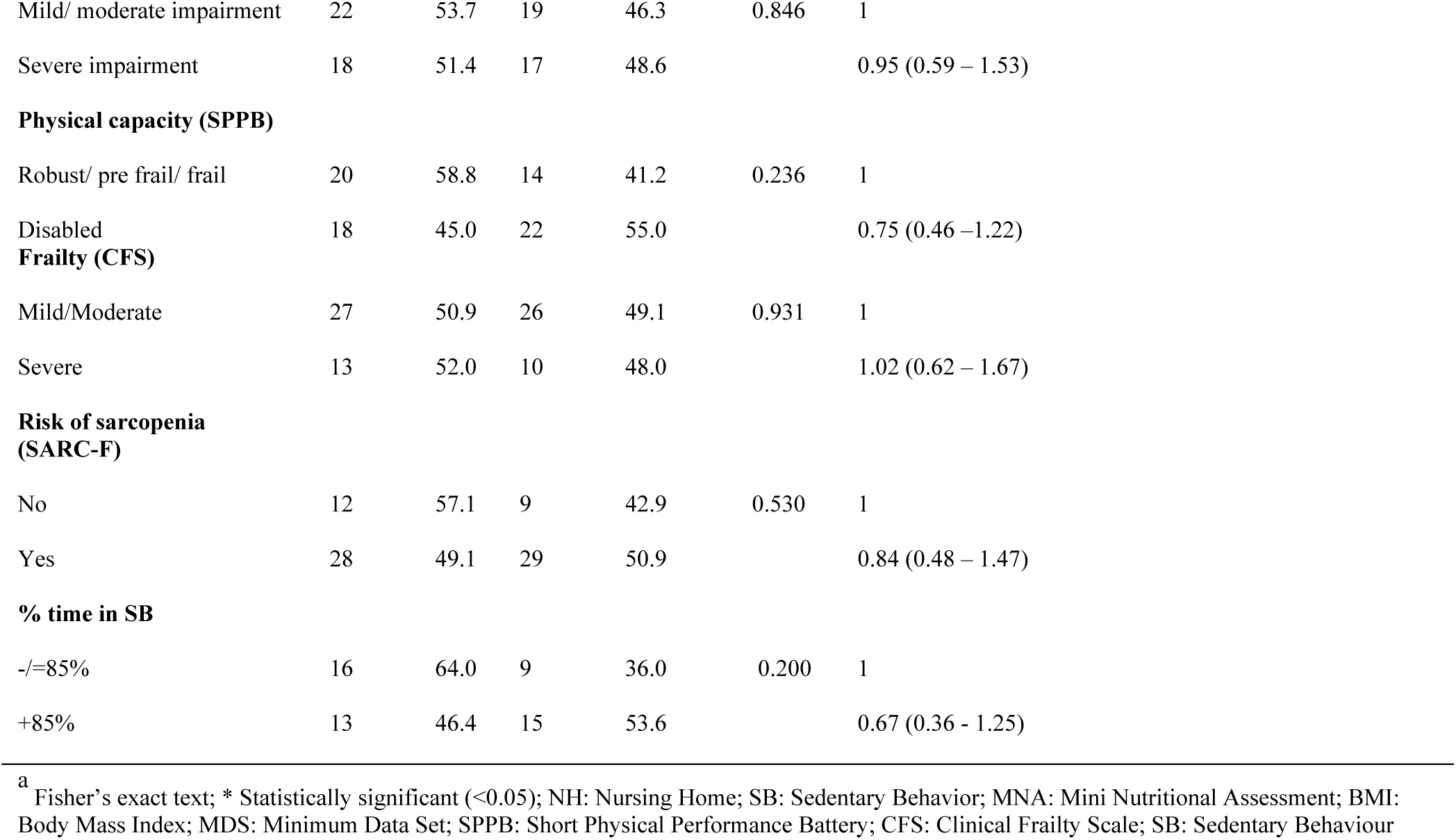
Bivariate and multivariate analyses showing the association of the variables and symptomatic COVID-19

| Symptomatic COVID-19 |  |  |  |  |  |  |  |  |
| --- | --- | --- | --- | --- | --- | --- | --- | --- |
| Variable | Yes (n=40) |  | No (n=38) |  | Bivariate analysis |  | Multivariate analysis |  |
|  | *n* | (%) | *n* | (%) | *p/Fisher* | RR (95% CI) | *p* | *Adjusted* RR (95% CI) |
| Age |  |  |  |  |  |  |  |  |
| -/≤ 85 years | 14 | 45.2 | 17 | 54.8 | 0.380 | 1 | 0.991 | 1.00 (0.92-1.10) |
| + 86 years | 26 | 55.3 | 21 | 44.7 |  | 1.22 (0.78– 1.92) |  |  |
| Sex |  |  |  |  |  |  |  |  |
| Men | 10 | 62.5 | 6 | 37.5 | 0.314 | 1 | 0.389 | 0.57 (0.16-2.01) |
| Women | 30 | 48.4 | 32 | 51.6 |  | 0.73 (0.37 – 1.43) |  |  |
| Marital status |  |  |  |  |  |  |  |  |
| Married/Partner | 5 | 41.7 | 7 | 58.3 | 0.636 | 1 |  |  |
| Single/Widowed | 29 | 49.2 | 30 | 50.8 |  | 1.14 (0.67 – 1.97) |  |  |
| Children |  |  |  |  |  |  |  |  |
| 0 – 1 | 16 | 43.2 | 21 | 56.8 | 0.177 | 1 |  |  |
| 2+ | 24 | 58.5 | 17 | 41.5 |  | 1.37 (0.86 – 2.16) |  |  |
| NH type |  |  |  |  |  |  |  |  |
| State subsidized places | 21 | 40.4 | 31 | 59.6 | **0.008\*** | 1 | **0.023\*** | 3.64 (1.20-11.04) |
| Private | 19 | 73.1 | 7 | 26.9 |  | 1.54 (1.11 – 2.15) |  |  |
| **Chronic diseases** |  |  |  |  |  |  |  |  |
| - 5 diseases | 15 | 42.9 | 20 | 57.1 | 0.177 | 1 |  |  |
| 5/ + diseases | 25 | 58.1 | 18 | 41.9 |  | 1.36 (0.86 – 2.15) |  |  |
| **Stroke** |  |  |  |  |  |  |  |  |
| No | 27 | 44.3 | 34 | 55.7 | **0.019\*** | 1 | **0.033\*** | 4.10 (1.12-14.72) |
| Yes | 13 | 76.5 | 4 | 23.5 |  | 2.36 (0.97 – 5.74) |  |  |
| **Smoke** |  |  |  |  |  |  |  |  |
| No | 27 | 51.9 | 25 | 48.1 | 1.000<sup>a</sup> | 1 |  |  |
| Yes | 2 | 50.0 | 2 | 50.0 |  | 0.96 (0.34 – 2.66) |  |  |
| **Alcohol** |  |  |  |  |  |  |  |  |
| No drinker | 26 | 45.6 | 31 | 54.4 | 1.000<sup>a</sup> | 1 |  |  |
| Drinker | 4 | 44.4 | 5 | 55.6 |  | 0.98 (0.52 – 1.84) |  |  |
| **Hospitalizations** |  |  |  |  |  |  |  |  |
| No | 33 | 53.2 | 29 | 46.8 | 0.499 | 1 |  |  |
| Yes | 7 | 43.8 | 9 | 56.3 |  | 0.83 (0.50 – 1.38) |  |  |
| **Nutritional Status (MNA)** |  |  |  |  |  |  |  |  |
| Normal | 8 | 42.1 | 11 | 57.9 | 0.490 | 1 |  |  |
| Malnutrition or risk | 8 | 32.0 | 17 | 68.0 |  | 0.85 (0.53 – 1.36) |  |  |
| **Weight loss (MNA)** |  |  |  |  |  |  |  |  |
| No | 35 | 53.8 | 30 | 46.2 | 0.606 | 1 |  |  |
| Yes | 5 | 45.5 | 7 | 54.5 |  | 0.86 (0.47 – 1.56) |  |  |
| **Obesity (BMI)** |  |  |  |  |  |  |  |  |
| No (- 30 BMI) | 6 | 42.9 | 8 | 57.1 | 0.744 | 1 |  |  |
| Yes (=/+ 30 BMI) | 22 | 47.8 | 24 | 52.2 |  | 1.09 (0.64 – 1.86) |  |  |
| **Urinary incontinence (MDS)** |  |  |  |  |  |  |  |  |
| No | 18 | 64.3 | 10 | 35.7 | 0.086 | 1 |  |  |
| Yes | 22 | 44.0 | 28 | 56.0 |  | 0.64 (0.36 – 1.11) |  |  |
| **Fecal incontinence (MDS)** |  |  |  |  |  |  |  |  |
| No | 33 | 54.1 | 28 | 45.9 | 0.448 | 1 |  |  |
| Yes | 6 | 42.9 | 8 | 57.1 |  | 0.80 (0.47 – 1.36) |  |  |
| **Functional capacity (Barthel Index)** |  |  |  |  |  |  |  |  |
| Independence/ slight/ moderate dependence | 26 | 57.8 | 19 | 42.2 | 0.180 | 1 |  |  |
| Severe dependence | 14 | 42.4 | 19 | 57.6 |  | 0.73 (0.46 – 1.15) |  |  |
| **Cognitive status (Pfeiffer Scale)** |  |  |  |  |  |  |  |  |

|  |  |  |  |  |  |  |
| --- | --- | --- | --- | --- | --- | --- |
| Mild/ moderate impairment | 22 | 53.7 | 19 | 46.3 | 0.846 | 1 |
| Severe impairment | 18 | 51.4 | 17 | 48.6 |  | 0.95 (0.59 – 1.53) |
| **Physical capacity (SPPB)** |  |  |  |  |  |  |
| Robust/ pre frail/ frail | 20 | 58.8 | 14 | 41.2 | 0.236 | 1 |
| Disabled | 18 | 45.0 | 22 | 55.0 |  | 0.75 (0.46 – 1.22) |
| **Frailty (CFS)** |  |  |  |  |  |  |
| Mild/Moderate | 27 | 50.9 | 26 | 49.1 | 0.931 | 1 |
| Severe | 13 | 52.0 | 10 | 48.0 |  | 1.02 (0.62 – 1.67) |
| **Risk of sarcopenia (SARC-F)** |  |  |  |  |  |  |
| No | 12 | 57.1 | 9 | 42.9 | 0.530 | 1 |
| Yes | 28 | 49.1 | 29 | 50.9 |  | 0.84 (0.48 – 1.47) |
| **% time in SB** |  |  |  |  |  |  |
| -/=85% | 16 | 64.0 | 9 | 36.0 | 0.200 | 1 |
| +85% | 13 | 46.4 | 15 | 53.6 |  | 0.67 (0.36 – 1.25) |

<sup>a</sup> Fisher's exact test; \* Statistically significant (<0.05); NH: Nursing Home; SB: Sedentary Behavior; MNA: Mini Nutritional Assessment; BMI: Body Mass Index; MDS: Minimum Data Set; SPPB: Short Physical Performance Battery; CFS: Clinical Frailty Scale; SB: Sedentary Behaviour

## DISCUSSION

The emergence of COVID-19 represents a dynamic, specific and real threat to the health and wellbeing of older people. The main objective of this study was to identify whether the health and sociodemographic conditions of institutionalized older people could influence having COVID-19 infection with symptoms.

In relation to our first finding, only half of the infected NH residents had COVID-19 symptoms, even with a confirmed diagnosis of COVID-19 infection. International evidence has shown that people living in NHs are particularly vulnerable to severe COVID-19 infections, nevertheless, the non-specific presentation of COVID-19 illness in older people has been highlighted by some scientific societies^24^. International comparisons of the data available are difficult due to differences in testing capabilities and policies (particularly in the earlier part of the pandemic), different approaches to recording deaths, and differing definitions of what constitutes a NH^25^.

In this sample, it was observed that having suffered a stroke and living in a private NH were positively associated with having symptomatic COVID-19, independently of age and sex. Previous studies have shown that older people with chronic conditions, including cerebrovascular pathology, had a higher risk of contracting COVID-19 infection with symptoms and consequently a higher risk of mortality from COVID-19^26,27^. However, the causative relationship between cerebrovascular events and COVID-19 is not conclusive^27^. Neurological pathologies have a component that compromises autoimmune aggression and, therefore, leads to the need for immunosuppressive or immunomodulatory management^28^. Inflammation occurs due to excessive activity of the immune system, derived from antigenic epitopes and pro-inflammatory molecules, in addition to the use of therapies that trigger the regulation of immune cells, causing in some cases affecting the innate and adaptive immunity of the person, leaving an altered immune system^29^. On the other hand, the virus has the ability to invade the brain parenchyma, endothelium, heart and alter clotting, and as a result is weaker immune function as a result of stroke, so the symptoms that manifest in these patients are major^30,31^.

Previous litearture^32^ have shown that the type and mainly the size of NHs (measured in the number of resident beds) influences the incidence of COVID-19 infection within NHs as in our study. The results of our study also identified that the type of residence, in this case private, could be related to a greater probability of suffering symptomatic COVID-19, with a greater risk of severity and complications. Braun et al., (2020) attributed these results to the shortage of PPE in private NHs^33^. Around 75% of NHs in Spain are private, with around 271696 private places, compared to 101289 publicly owned^34^. Studies have shown that the larger the size of the NH, the higher the risk of death from COVID-19^35^. Inadequate infrastructure, lack of space and high occupancy of NHs at the time of the crisis increased the chances of contracting COVID-19 with more symptomatology^9^. It is important to identify those factors that are modifiable and found within the institution and the different levels of pathology transmission in NHs^36^.

Nursing homes have a disproportionately high level of risk for COVID-19 transmission^37^, for this reason, recording COVID-19 mortality in a NH can inform aspects of risk management and allows for optimizing care in future interventions in NHs^36^.

The main limitation of this study is the small sample size; nevertheless, the pandemic outbreak in March 2020 marked the end of the baseline NHs data collection. In addition, we are uncertain about the accuracy of serologic testing in persons with mild or no symptoms^37^, but this study was based on the tools used in the NHs and the existing literature at the time.

In terms of implications for clinical practice, it is important to screen institutionalized older adults who suffered a stroke since they are at increased risk for symptomatic COVID-19 in order to provide early care. Now we know more about COVID-19 and its impact on NHs, social and health services should improve and tailor contingency plans to prevent the spread of infectious diseases in NHs to protect the most vulnerable and frail population segment of our society. Special attention to address environmental factors such as inadequate infrastructure, poorly ventilated environment, high occupancy and low staffing is required.

## CONCLUSIONS

It can be concluded that having suffered a stroke and living permanently in a private NH represented risk factors for symptomatic COVID-19 in this sample of institutionalized older people. A large, adequately powered study is recommended to support the results from this research.

## Data Availability

All relevant data are within the manuscript and its Supporting Information files.

## Acknowledgements

We thank all the members of the study NH for their contribution to this work and the older adults who participated in the study.

## REFERENCES

1. Licher S, Terzikhan N, Splinter MJ, et al. Design, implementation and initial findings of COVID-19 research in the Rotterdam Study: leveraging existing infrastructure for population-based investigations on an emerging disease. Eur J Epidemiol [Internet]. 2021;36(6):649–54. Available from: https://doi.org/10.1007/s10654-021-00789-7

2. Madabhavi I, Sarkar M, Kadakol N. CoviD-19: A review. Monaldi Arch Chest Dis. 2020;90(2):248–58.

3. Barrera-Algarín E. COVID-19 y personas mayores en residencias: impacto según el tipo de residencia. 2020;(January).

4. Dhama K, Patel SK, Kumar R, et al. Geriatric Population During the COVID-19 Pandemic: Problems, Considerations, Exigencies, and Beyond. Front Public Heal. 2020;8(September):1–8.

5. Davis P, Gibson R, Wright E, et al. Atypical presentations in the hospitalised older adult testing positive for SARS-CoV-2: a retrospective observational study in Glasgow, Scotland. Scott Med J. 2021;66(2):89–97.

6. Mumm J, Osterman A, Ruzicka M, et al. Urinary Frequency as a Possibly Overlooked Symptom in COVID-19 Patients: Does SARS-CoV-2 Cause Viral Cystitis? 2020;(January).

7. Cipriani G, Danti S, Nuti A, Carlesi C, Lucetti C, Di Fiorino M. A complication of coronavirus disease 2019: delirium. Acta Neurol Belg [Internet]. 2020;120(4):927–32. Available from: https://doi.org/10.1007/s13760-020-01401-7.

8. España PP, Bilbao A, García-Gutiérrez S, et al. Predictors of mortality of COVID-19 in the general population and nursing homes. Intern Emerg Med. 2021 Sep;16(6):1487–96.

9. Barrera-Algarín E, Estepa-Maestre F, Sarasola-Sánchez-Serrano JL, Malagón-Siria JC. COVID-19 y personas mayores en residencias: impacto según el tipo de residencia. Rev Esp Geriatr Gerontol. 2021 Jul;56(4):208–17.

10. Fité-Serra AM, Gea-Sánchez M, Alconada-Romero Á, et al. Occupational precariousness of nursing staff in catalonia’s public and private nursing homes. Int J Environ Res Public Health. 2019 Dec;16(24).

11. Navarro Prados AB, Jiménez García-Tizón S, Meléndez JC. Sense of coherence and burnout in nursing home workers during the COVID-19 pandemic in Spain. Heal Soc Care Community. 2021;(July 2020):1–9.

12. von Elm E, Altman DG, Egger M, Pocock SJ, Gøtzsche PC, Vandenbroucke JP. The Strengthening the Reporting of Observational Studies in Epidemiology (STROBE) statement: guidelines for reporting observational studies. J Clin Epidemiol. 2008;61(4):344–9.

13. Farrés-Godayol P, Jerez-Roig J, Minobes-Molina E, et al. Urinary incontinence and sedentary behaviour in nursing home residents in Osona, Catalonia: protocol for the OsoNaH project, a multicentre observational study. BMJ Open [Internet]. 2021;11(4):e041152. Available from: http://www.ncbi.nlm.nih.gov/pubmed/33879481

14. López de la Iglesia J, Fernández-Villa T, Rivero A, et al. Factores predictivos de COVID-19 en pacientes conRT-qPCR negativo. 2020;(January).

15. Dewar AM, Thornhill WA, Read LA. The effects of tefluthrin on beneficial insects in sugar beet. Mini Nutr Assess Its Use Grading Nutr State Elder Patients. 1994;15(3):987–92.

16. Klusch L. The MDS 3.0 and its impact on bladder and bowel care. Provider. 2012;38(6):33, 35, 37 passim.

17. Baztán JJ. Índice de Barthel: Instrumento válido para la valoración funcional de pacientes con enfermedad cerebrovascular. 2016;(March).

18. Martinez de la Iglesia J, Duenas Herrero R, Onis Vilches MC, Aguado Taberne C, Albert Colomer C, Luque Luque R. Spanish language adaptation and validation of the Pfeiffer’s questionnaire (SPMSQ) to detect cognitive deterioration in people over 65 years of age. Med Clin (Barc). 2001;117(4):129– 34.

19. Malmstrom TK, Miller DK, Simonsick EM, Ferrucci L, Morley JE. SARC-F: A symptom score to predict persons with sarcopenia at risk for poor functional outcomes. J Cachexia Sarcopenia Muscle. 2016;7(1):28–36.

20. Edwardson CL, Rowlands AV., Bunnewell S, et al. Accuracy of posture allocation algorithms for thigh- and waist-worn accelerometers. Med Sci Sports Exerc. 2016;48(6):1085–90.

21. Geidl W, Knocke K, Schupp W, Pfeifer K. Measuring stroke patients’ exercise preferences using a discrete choice experiment. Neurol Int. 2018;10(1):13–7.

22. Rockwood K, Song X, MacKnight C et al. A global clinical measure of fitness and frailty in elderly people. Cmaj. 2005;173(5):489–95.

23. Cabrero-García J, Muñoz-Mendoza CL, Cabañero-Martínez MJ, González-Llopís L, Ramos-Pichardo JD, Reig-Ferrer A. Valores de referencia de la Short Physical Performance Battery para pacientes de 70 y más años en atención primaria de salud. Aten Primaria. 2012;44(9):540–8.

24. Lithander FE, Neumann S, Tenison E, et al. COVID-19 in older people: A rapid clinical review. Age Ageing. 2020;49(4):501–15.

25. Comas-Herrera A, Zalakain J. Mortality associated with COVID-19 outbreaks in care homes: early international evidence. Resour to Support community institutional Long-Term Care responses to COVID-19 [Internet]. 2020;(April):1–6. Available from: https://www.ontario.ca/page/how-ontario-is-responding-covid-19#section-1%0ALTCcovid.org.

26. Graham NSN, Junghans C, Downes R, et al. SARS-CoV-2 infection, clinical features and outcome of COVID-19 in United Kingdom nursing homes. J Infect. 2020;81(3):411–9.

27. Wang Z, Yang Y, Liang X, et al. COVID-19 Associated Ischemic Stroke and Hemorrhagic Stroke: Incidence, Potential Pathological Mechanism, and Management. Front Neurol. 2020;11(October):1–8.

28. Bossù P, Toppi E, Sterbini V, Spalletta G. Implication of aging related chronic neuroinflammation on covid-19 pandemic. J Pers Med. 2020;10(3):1–13.

29. Sonja A. Rasmussen, MD, Ms JCS. Since January 2020 Elsevier has created a COVID-19 resource centre with free information in English and Mandarin on the novel coronavirus COVID-. Ann Oncol. 2020;(January):19–21.

30. Trejo-Gabriel-Galán JM. Stroke as a complication and prognostic factor of COVID-19. Neurologia. 2020;35(5):318–22.

31. Chen N, Zhou M, Dong X, et al. Epidemiological and Clinical Characteristics of 99 Cases of 2019 Novel Coronavirus Pneumonia in Wuhan, China: A Descript. Lancet. 2020;395(10223):507–13.

32. Burton JK, Bayne G, Evans C, et al. Evolution and effects of COVID-19 outbreaks in care homes: a population analysis in 189 care homes in one geographical region of the UK. 2020;(January):19–21.

33. Braun RT, Yun H, Casalino LP, et al. Comparative Performance of Private Equity-Owned US Nursing Homes during the COVID-19 Pandemic. JAMA Netw Open. 2020;3(10):1–11.

34. Abellán A, Aceituno P, Pérez J, Ramiro D, Ayala A, Puyol R. Informes Envejecimiento en red. Indicadores estadísticos básicos. 2019;(March):38.

35. Zimmerman S, Edd CD, Tandan M, et al. Nontraditional Small House Nursing Homes Have Fewer COVID-19 Cases and Deaths. 2020;(January).

36. Ibrahim JE, Li Y, McKee G, Eren H, et al. Characteristics of nursing homes associated with COVID-19 outbreaks and mortality among residents in Victoria, Australia. Australas J Ageing. 2021;(May):1–10.

37. Chen M, Qin R, Jiang M, et al. Clinical applications of detecting IgG, IgM or IgA antibody for the diagnosis of COVID-19: A meta-analysis and systematic review. 2020;(January).

